# Pediatric human immunodeficiency virus-positive disclosure status and associated factors among caregivers of children in Wolaita and Hadiya zone, southern Ethiopia

**DOI:** 10.1101/2024.12.03.24318443

**Authors:** Lina Tesfaye, Amene Abebe, Simegn Molla, Amare Admasu

**Author notes:** **Authors’ contribution** All authors contributed equally.

## Abstract

**Background:** Human immunodeficiency virus (HIV) positive status disclosure is an essential component of pediatric care and long-term disease management. However, pediatric HIV disclosure is a complex and understudied public health concern. This study aimed to assess the pediatric HIV-positive disclosure status and associated factors among caregivers.

**Method:** A facility-based cross-sectional study was done among 375 caregivers of HIV-positive children in selected facilities from March 1 to April 30, 2022. Data was collected through inperson interviews using a carefully designed questionnaire that had been tested beforehand. Study participants were randomly selected from the anti-retroviral therapy (ART) logbook using a lottery method as the sampling frame. The data collected was inputted into Epidata version 3.02 and then transferred to SPSS version 23 for analysis. The findings were displayed through tables, graphs, charts, and written descriptions. Statistical analyses using different models were performed to examine the data. The association of variables was declared at 95%CI and p-value <0.05 and the strength of association was determined using the adjusted odds ratio (AOR).

**Result:** Out of 371 participants, 98.9% replied to the survey. Forty-one point eight percent of caregivers told them about their HIV-positive children’s status. Caregivers/parents’ discussions with health care providers about disclosure [AOR 2.171, 95% CI [1.199, 3.931]], child duration on ART [AOR 1.633, 95% CI [1.013, 2.631]], and child stigmatization [AOR 2.103, 95% CI [1.361, 3.250]] were significantly associated with pediatric HIV positive status disclosure.

**Conclusion:** The rate of disclosing pediatric HIV-positive status was lower compared to other studies in Ethiopia. Caregivers who talked to healthcare providers, children on ART for a longer time, and caregivers not fearing stigma for their child were more likely to disclose pediatric HIV-positive status. Facility management and healthcare providers should focus on improving the disclosure of pediatric HIV-positive status.

## Introduction

### Background

Disclosure refers to when a child is aware of their HIV status with evidence supporting the benefits of suitable pediatric care (1). The advantages of disclosure involve increased adherence to treatment, better healthcare outcomes, and improved communication between young individuals, caregivers, and medical professionals (2). However, revealing a child’s HIV-positive status is contentious and emotionally challenging for caregivers due to concerns about the child maintaining confidentiality, potential disclosure of family secrets, and fear of community stigma leading to social repercussions like rejection by healthcare providers, parents, or caregivers (3).

HIV/AIDS is increasingly affecting the health of children and undermining their survival in highly affected countries (4). The WHO reported around 12,146 deaths related to AIDS in 2019 (5). According to a 2007 report review, disclosure rates in North American and European studies varied from 10% to 75% (6). A systematic review found that HIV-positive status disclosure to HIV-infected children in low-and middle-income countries was generally low, ranging from 1.7% to 41% (7).

Children with HIV illness have been dubbed “the missing face of AIDS” because, unlike adults, they frequently lack basic health care and have been “missing from global and national policy discussions” (8). However, the benefits extend beyond the kid; they also help caregivers by easing the stress and anxiety associated with secrecy and deceit, as well as offering a chance to create a trusting connection and more open communication with the children (1). They also obtained spiritual and emotional fulfillment from drug-induced despondency and opportunistic infection, as well as support from their family over a question. In contrast, in resource-constrained contexts, recorded findings found relatively low levels of disclosure (3). Research from low-and middle-income nations showed that the amount of HIV status disclosure ranges from 0–69%. In five sub-Saharan African countries, a concerning 67.3% of caregivers chose not to reveal their HIV status to their children, highlighting the challenges of disclosure in this region (9).

The African Network for the Care of Children Affected by HIV/AIDS (ANECCA) also recommends that informing children about their HIV status should start as early as 5–7 years old (10). Similarly, Ethiopian national HIV/AIDS treatment guidelines recommend that children should be informed about their HIV status starting as early as 4–6 years of age and gradually over time (11). According to Ethiopian studies, as cited by Tucho WA, Tekelehaimanot AN, and Habte MB., 2021, “the proportion of children who are HIV-positive ranges from 28.5 to 49.4%, as stated by Tadesse BT et al., 2015; Mengesha MM et al., 2018; Abegaz BF et al., 2019; and Lencha B., 2018” (12). Many studies show that informing children about their HIV status allows them to seek social support without the risk of accidental disclosure and may reduce disruptive behavior. It would also help to ensure that children take their medications as prescribed, develop coping skills, and practice safer sex to prevent the spread of HIV (1). Therefore, this study aimed to investigate informing children about their HIV status and related factors among HIV-infected children on ART.

Fewer studies, conducted in various Ethiopian cities, revealed that the percentage of children who knew they were HIV positive ranged from 16.3% to 49% (4). Lack of disclosure may lead to accidental disclosure from overhearing caregiver discussions. This, in turn, may lead to both maladjustment and distrust of adults (13). There are many barriers to timely disclosure of HIV-positive status to children. These include how the child reacts to the news of the diagnosis, the age of the child, the lack of caregiver readiness to discuss how the child got infected, and the fear of the child about keeping the status a secret (10).

The outcomes of this study will shed light on the factors influencing pediatric HIV-positive status disclosure. This study’s findings may aid in the development of initiatives to improve the disclosure of HIV status to youngsters. In addition, this study will detect disparities among parents/caregivers in revealing HIV status to children. This information will help hospital administrators, woreda health offices, and health facilities develop specialized plans and strategies to successfully manage disclosure concerns. This study will provide fundamental data for hospital management bodies, woreda health offices, health center directors, program managers, and implementers at all levels to address gaps in HIV-positive status disclosure to children living with HIV. It may encourage other scholars and policymakers to perform further study in this area.

## Method and materials

The study was conducted in health facilities in Hadiya and Wolaita Zone. Wolaita Zone, which is located 380 kilometers south of Addis Ababa, and Hadiya Zone from March 1 to April 30, 2022. Wolaita Zone, located 380 kilometers south of Addis Ababa, has a total of 4,210 people who are HIV/AIDS-positive, including 160 children. Twenty health facilities, including hospitals and health centers, provide ART services in the Wolaita Zone.

Hadiya Zone, located 280 kilometers south of Addis Abeba, has a total population of 3,970 HIV/ADIS-positive persons, including 270 children. In the Hadiya zone, seventeen health institutions, including hospitals and health centers, provide antiretroviral therapy (ART). In both zones, ART unit services are delivered by a team that includes the focal person, case manager, adherence supporter, health care practitioners, and data clerk personnel. Additionally, pediatric psychosocial training sessions are held at the Dubo hospital in the Wolaita zone, as well as the Shone primary hospital and the Nigist Eleni Mohamed Memorial hospital in the Hadiya zone.

A facility-based cross-sectional study was conducted, with quantitative data collected. From all HIV-positive children aged 6 to 15 and their caregivers in selected health facilities in Wolaita and Hadiya zones. The study included caregivers of HIV-positive children aged 6 to 15 who received services at the selected healthcare facilities during the study period.

### Inclusion criteria

Selected caregivers of HIV-positive pediatrics aged 6–15 on ART for the last 6 months.

### Exclusion criteria

Caregivers with other serious illnesses were excluded during the study period.

### Sample size determination

The sample size was determined by using a single population proportion formula and considering the following assumptions: 95% CI, 5% margin of error, and magnitude of HIV-positive status disclosure in southern Ethiopia (p), which is 33.3% (3)

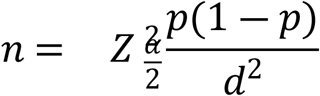

n=required sample size, Z= 1.96 at 95% confidence interval

d= margin of error, p = population proportion

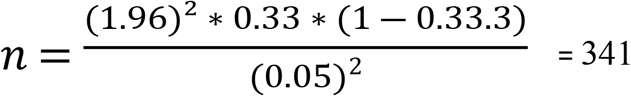

By adding a non-response rate of 10% = 375

### Sampling procedure

The study was carried out in randomly selected health facilities in the Wolaita and Hadiya zones. Twenty-two health centers and 11 hospitals were randomly chosen from all the health facilities providing ART services in both zones using a simple random sampling method. The sample size for the selected health facilities in both zones was determined based on the number of pediatric ART beneficiaries, with an allocation proportional to this count. A lottery method was employed to recruit 375 study participants, with 160 from health centers and 215 from hospitals. This selection was based on the client registration logbook, excluding drop-outs, transfer-outs, and individuals lost to follow-up from the randomly chosen hospitals and health centers (refer to Figure 1).

**Figure 1.**
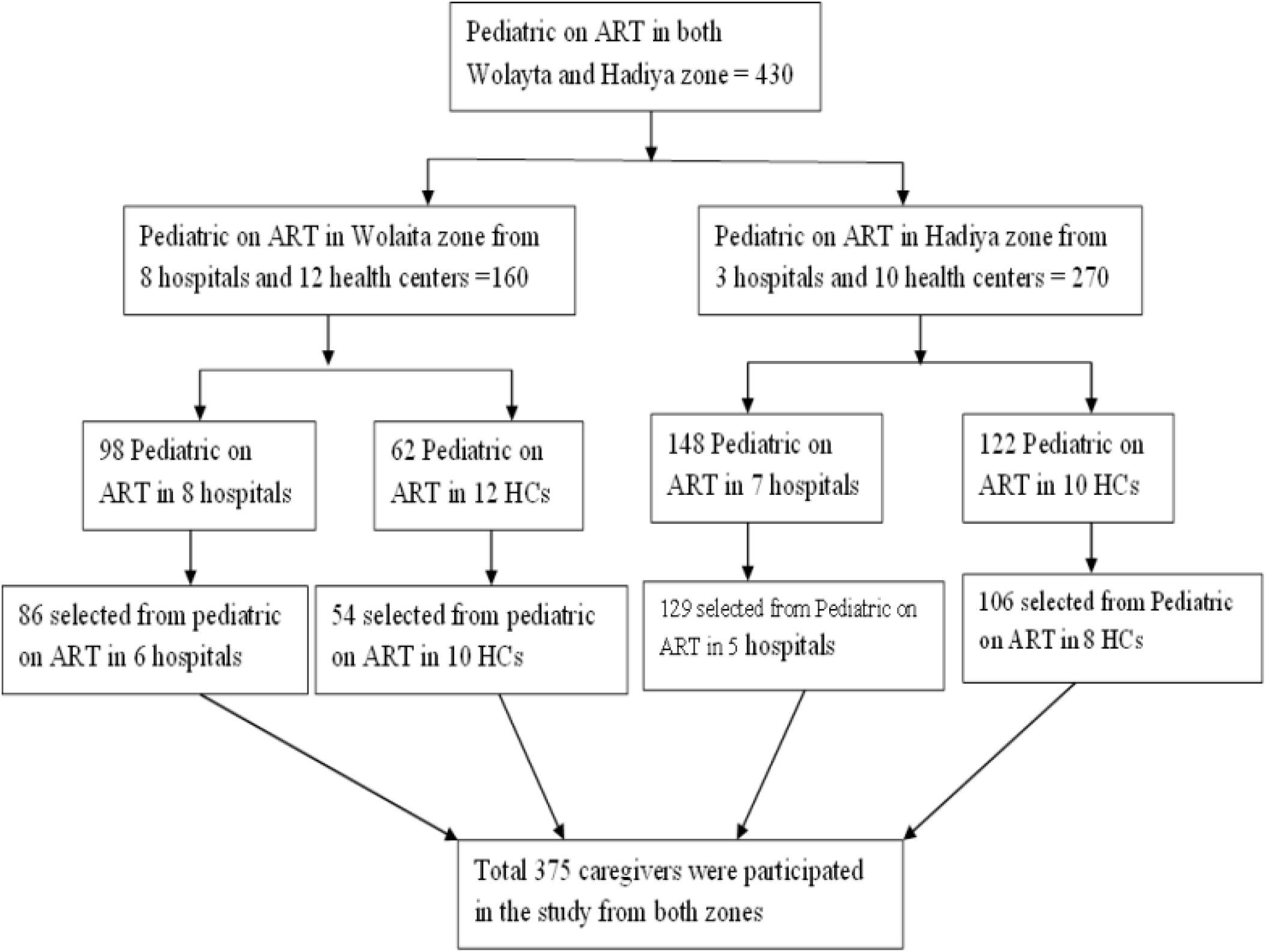
schematic presentation of sampling techniques

### Dependent variable

Pediatric HIV positive disclosure Status

### Independent variables

Factors to consider include the child’s age, gender, loss of family members, and religion. Medical aspects to consider include age at commencement of antiretroviral therapy, length of antiretroviral therapy, age at HIV diagnosis, antiretroviral therapy follow-up location, treatment support availability, the child’s responsibility for antiretroviral therapy, and the right age for disclosure. Factors to consider: social support, stigma, and discrimination. Factors to examine include the caregiver’s understanding of HIV disclosure, their education level, discussions between health experts and parents about disclosure, and the caregiver’s connection with the kid.

### Operational definition

- HIV positive disclosure: The act of informing HIV positive status (17)
- Full disclosure: was generally considered as “full” when it involved the caregiver having disclosed to the child that he or she has HIV(8).
- *Pediatrics: the child has HIV between the 6-15 age groups (14)*.
- Caregiver: the person who lives with HIV positive child and knows about the child’s HIV status (17).
- Adherence: attends to their regular clinical follow-up care, and periodic laboratory monitoring, and avoids practices that interfere with treatment effectiveness (3).

### Data collection tool and method

Data were collected using semi-structured, pretested questionnaires. The questionnaire consists of four parts. Part I: Caregiver Part II of the socio-demographic questionnaire for children. Part III contains questions concerning clinical issues, whereas Part IV contains questions about psychosocial aspects (disclosure-related).

### Data collection procedure

Data collection was carried out by ART-trained nurses or health officials. Initially, an English-language questionnaire was produced. The English version of the questionnaire was translated into the local languages of Wolaitato and Hadiya before being back-translated to English by language specialists to ensure its original meaning. Nurses or health officials acquired data through face-to-face interviews with pre-tested semi-structured questionnaires. Six nurses and three health officers were hired in the zone to gather data and supervise patients. During the children’s follow-up visits to the ART clinic, their caregivers were questioned in a separate room.

### Data quality assurance

Data quality was controlled through intensive training of data collectors on objectives, the questionnaire, and the method of data collection for one day. Data quality was ensured during collection, coding, entry, and analysis. Data quality was also controlled by conducting a pre-test on 5% of the sample at Kulito and Halaba to assess clarity, understanding ability, flow, and consistency. The test was revised before the start of data collection, and important modifications were made based on the findings. The entire questionnaire was supervised and reviewed for completeness and consistency. The completeness of the questionnaire was also checked before data entry.

### Data processing and analysis

The data were entered into Epi Data Version 3.1 for data collection and then exported to SPSS 25 for cleaning and analysis. Descriptive statistics were computed to summarize and analyze the data. Statistics such as frequencies, percentages, and means were used to analyze the data. In a multivariable logistic regression model, all variables with a p-value of 0.25 in the n-variable analysis were entered to control for all possible confounders and identify factors of the outcome variable. An adjusted odds ratio and a 95% confidence interval were estimated to assess the strength of the association between variables. Multicollinearity was detected by checking the VIF (variance inflation factor), which assesses the correlation between independent variables. The model’s fitness was evaluated using the Hosmer-Lemeshow goodness-of-fit test to assess how well it fits the observed data. Statistical significance was determined at a p-value of 0.05, indicating the level at which results are considered statistically significant. The outcome is presented via text, table, and graph.

### Ethical considerations

The Wolaita Sodo University Research and Institutional Review Board (RIRB) provided ethical clearance for the project. Both zones received an approval from the RIRB granting them access to the health facilities. Consent was gained from parents or legal guardians after notifying them of the study’s purpose. All participants were given secrecy and the freedom to decline. To maintain anonymity, no personal identifiable information was collected, ensuring confidentiality and privacy.

## Result

### Socio-demographic characteristics of respondents/caregivers

The study included 371 caregivers from the predicted sample of 375. This yielded a response rate of 98.9%. Out of 371 participants in the research, 220 (59.3%) were women. The average age of respondents was 37.82 years, with a standard deviation of 8.4, indicating a range of ages around the norm. In terms of religion, 199 (53.6%) were protestants, with the bulk of 217 (58.5%) coming from cities. In terms of caregiver relationships, more than half (217, or 58.5%) were the children’s biological parents, whereas one-third were farmers. See Table 1 for further information.

**Table 1:** Socio-demographic characteristics of respondents/caregivers in Wolaita and Hadiya Zone, Southern Ethiopia, 2022.

| **Variables** | **Category** | **Frequency** | **Percent** |
| --- | --- | --- | --- |
| **Age of caregivers** | <30 years | 68 | 18.3 |
|  | 31-40 years | 215 | 58 |
|  | >40 years | 88 | 23.7 |
| **Ethnicity of caregivers** | Wolaita | 159 | 42.9 |
|  | Hadiya | 152 | 41 |
|  | Gurage | 38 | 10.2 |
|  | Other | 22 | 5.9 |
| **Religion of caregivers** | Protestant | 199 | 53.6 |
|  | Orthodox | 147 | 39.6 |
|  | Muslim | 25 | 6.7 |
| **Marital status** | Single | 72 | 19.4 |
|  | Married | 210 | 56.6 |
|  | Divorced | 44 | 11.9 |
|  | Widowed | 25 | 6.7 |
|  | Separate | 20 | 5.4 |
| **Educational status** | No formal education | 167 | 45 |
|  | Elementary school | 132 | 35.6 |
|  | Secondary school | 53 | 14.3 |
|  | Collage and above | 19 | 5.1 |
| **Occupation** | Government worker | 99 | 26.7 |
|  | Farmer | 137 | 36.9 |
|  | Merchant | 99 | 26.7 |
|  | Other | 36 | 9.7 |
| **Monthly income in ETB** | <1000 | 118 | 31.8 |
|  | 1001-3000 | 194 | 52.3 |
|  | ≥3001 | 59 | 15.9 |

**Table 2:** Socio-demographic characteristics of pediatrics in Wolaita and Hadiya Zone, Southern Ethiopia, 2022.

| Variables | Category | Frequency | Percent |
| --- | --- | --- | --- |
| **Age of child** | <10 years | 158 | 42.6 |
|  | ≥10 years | 213 | 57.4 |
| **Sex of child** | Male | 188 | 50.7 |
|  | Female | 183 | 49.3 |
| **Child collect drug** | Yes | 255 | 68.7 |
|  | No | 116 | 31.3 |
| **Place drug collected** | Hospital | 222 | 59.8 |
|  | Health center | 149 | 40.2 |
| **A child with whom currently living** | Biological parent | 203 | 54.7 |
|  | Siblings | 122 | 32.9 |
|  | Relatives | 27 | 7.3 |
|  | At orphanage camp | 19 | 5.1 |
| **The child lost any family members by HIV** | Yes | 158 | 42.6 |
|  | No | 213 | 57.4 |
| **Parent lost** | Father | 66 | 17.8 |
|  | Mother | 49 | 13.2 |
|  | Both mother and father | 43 | 11.6 |

### Socio-demographic characteristics of pediatrics

Almost half (188, or 50.7%) of the children were boys, with an average age of 11.19 ±2.75 SD years. The bulk of the youngsters, 192 (51.8%) and 266 (71.7%), started formal schooling before the age of 10. Among the children, 203 (54.7%) still lived with their biological family, while 158 (42.6%) had lost at least one biological parent.

### Clinical characteristics of pediatrics and caregivers

More than two-thirds of the caregivers (68.7%) were HIV positive. The majority (348, or 93.8%) of the pediatric patients were diagnosed before the age of five, with the average age of diagnosis being 2.2 years (SD = 1.78). The average age of pediatric patients on antiretroviral treatment (ART) was 7.66 years (SD 4.27), and more than three-quarters (283, or 76.3%) had WHO clinical stage I illness, with 158, or 42.6%, having previously been hospitalized to the hospital (Table 3).

**Table 3:** Clinical characteristics of children and caregivers of respondents/caregivers in Wolaita and Hadiya Zone, Southern Ethiopia, 2022.

| Variables | Category | Frequency | Percent |
| --- | --- | --- | --- |
| **Age of pediatrics at HIV diagnosis** | <5 years | 348 | 93.8 |
|  | ≥5 years | 23 | 6.2 |
| **Pediatric duration on ART** | <5 years | 254 | 68.5 |
|  | 6-15 years | 117 | 31.5 |
| **pediatrics responsible for ART** | Yes | 255 | 68.7 |
|  | No | 116 | 31.3 |
| Current WHO clinical stage | Stage I | 283 | 76.3 |
|  | Stage II | 88 | 23.7 |
|  | Stage III | 0 | 0 |
|  | Stage IV | 0 | 0 |
| Caregiver on ART | Yes | 255 | 68.7 |
|  | No | 116 | 31.3 |
| Recent viral load | Suppressed | 113 | 30.5 |
|  | Not suppressed | 258 | 69.5 |
| Current opportunistic infection | Yes | 219 | 59 |
|  | No | 152 | 41 |

**Table 4:** Participant’s psychosocial support-related characteristics in Wolaita and Hadiya Zone, Southern Ethiopia, 2022.

| Variables | Category | Frequency | Percent |
| --- | --- | --- | --- |
| psychosocial support provided for the child | Yes | 116 | 31.3 |
|  | No | 255 | 68.7 |
| Material support is given to children | Yes | 258 | 69.5 |
|  | No | 113 | 30.5 |
| Kind of organization gives support | Government organization | 188 | 50.7 |
|  | Nongovernment organization | 43 | 11.6 |
|  | Religious organization | 27 | 7.3 |
| Individuals giving support to children | ART focal | 261 | 70.4 |
|  | ART provider | 93 | 25.1 |
|  | Case manager | 17 | 4.6 |
| Respondents' belief of psychosocial education | To disclose the children about their status | 227 | 61.2 |
|  | To provide correct information about HIV and self-care needs | 82 | 22.1 |
|  | For mental health education | 40 | 10.8 |
|  | Personal hygiene consideration | 22 | 5.9 |
| The separated room given for psycho-social support | Yes | 140 | 37.7 |
|  | No | 231 | 62.3 |

### Disclosure status of HIV-positive pediatrics

From an overall sample of 371 respondents or caregivers, the prevalence of disclosure of HIV-positive status to HIV-infected children in Wolaita and Hadiya zones was 155 (41.8%), with a 95% CI of 37–47 (figure 2).

**Figure 2:**
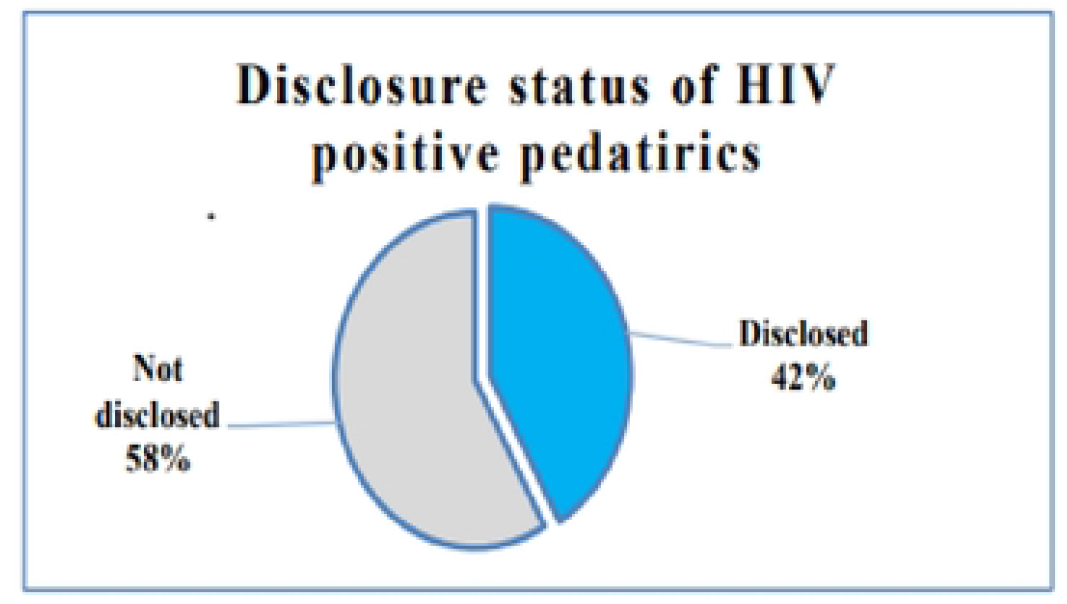
Participant’s disclosure HIV positive status to children living with HIV in Wolaita and Hadiya Zone, Southern Ethiopia, 2022

At the time of disclosure, the average age of the kid was 11.88 (SD±2.425) years. More than half of the 200 youngsters (53.9%) were aged 12 or older. More than half (56%) of the 155 caregivers who provided reasons for the disclosure, or 87 stated that the child was old enough (Figure 3).

**Figure 3:**
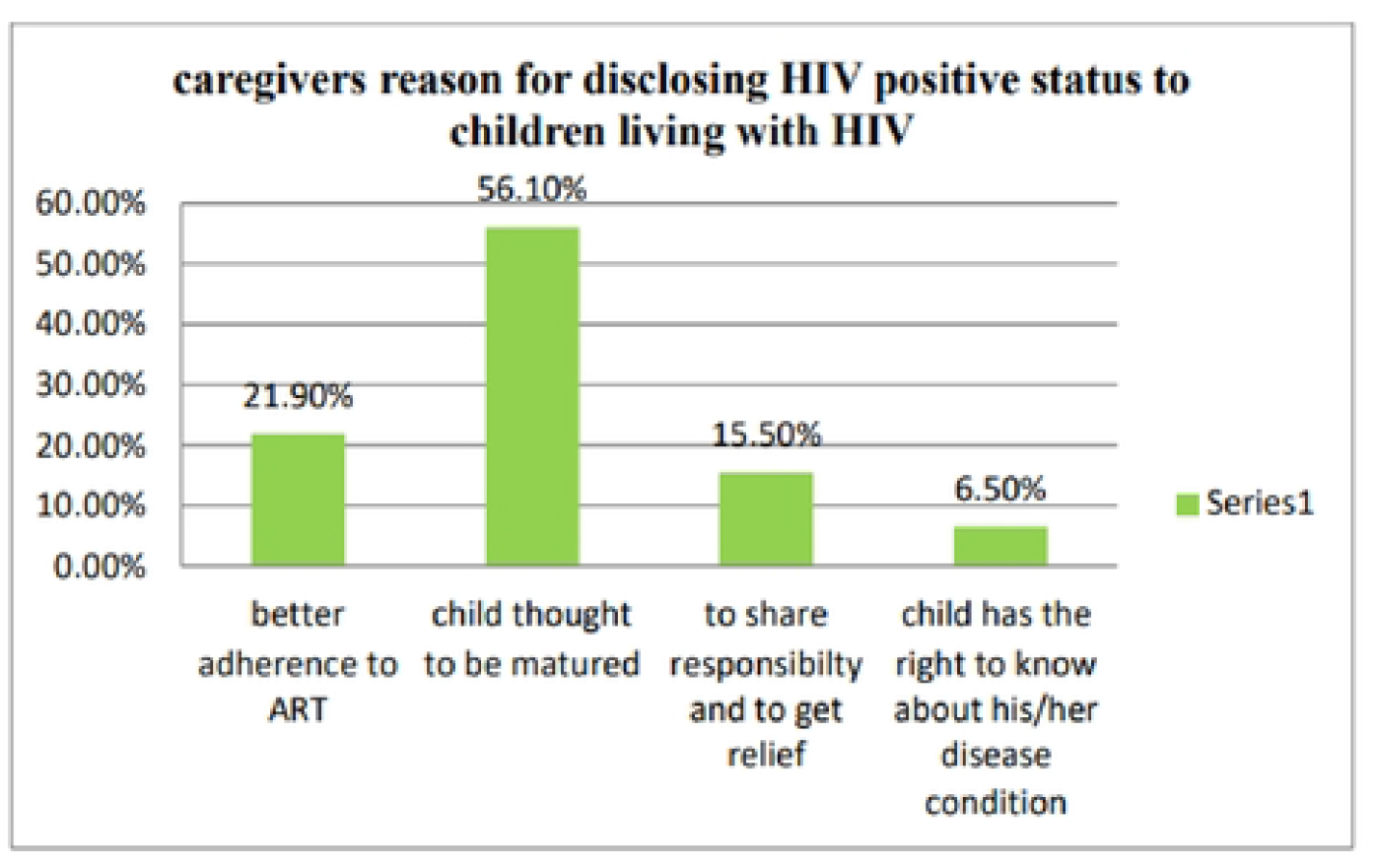
Participant’s reason for disclosure HIV positive status to children living with HIV in Wolaita and 1 ladiya Zone. Southern Ethiopia. 2022

The vast majority of 314 participants (84.6%) did not raise disclosure concerns with their healthcare providers. One hundred fifty-two (41%) of caregivers agreed that they should be responsible for disclosing pediatric HIV-positive status. One hundred eighty (48.5%) of caregivers said that both the health care practitioner and the caregiver should be accountable for disclosure. Thirty-nine (10.5%) caregivers agreed that the health care provider should be held accountable for giving disclosure. Furthermore, two-thirds (247, or 66.6%) of caregivers felt that revealing HIV status to HIV-positive children is a critical concern. More than half (53.9%) of caregivers believed that children above the age of 12 should reveal their HIV status to relevant people at the appropriate time. Caregivers stated that the child’s initial reaction after discovering their HIV status was tearful or crying (24.5%), angry and asking many questions (54.8%), and surprised with no reaction (20.6%).

In terms of why information is not provided, most caregivers (93.1%) believe the kid is open, while others are concerned that it would emotionally injure the child, lead to self-discrimination, and keep the child unconscious of the condition (Figure 4).

**Figure 4:**
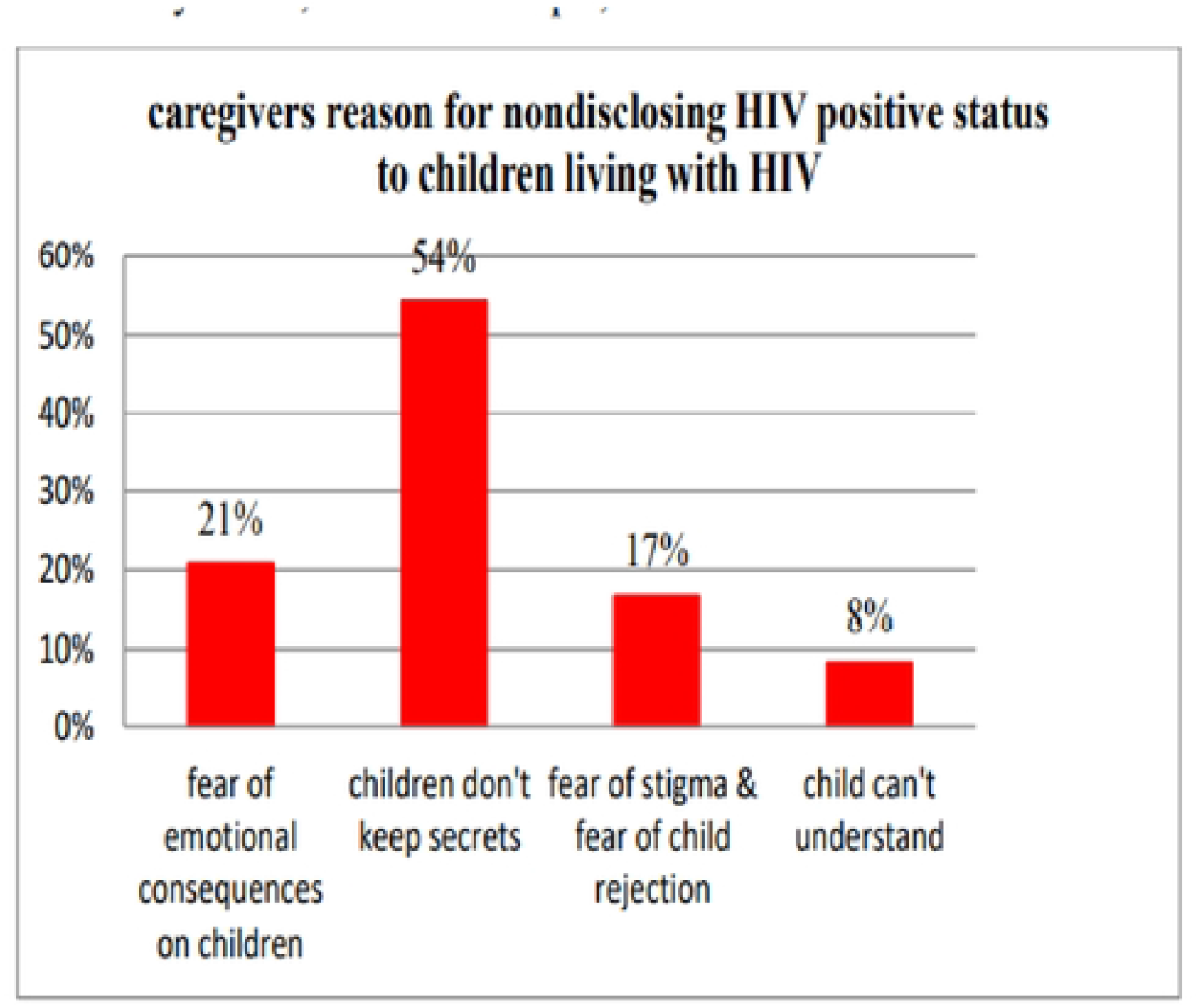
Participant’s reason for nondisclosure HIV positive status to children living with HIV in Wolaita and Hadiya Zone, Southern Ethiopia, 2022

### Relationship between disclosure and ART adherence

Out of 371 children, 304 (81.9%) had good treatment adherence, 12.4% were fair, and 5.7% were poor. Caregivers noted that not disclosing a child’s HIV status might result in medication cessation (43.4%), inefficient medicine intake by the kid (44.2%), and difficulty explaining the necessity for medication to family members (12.4%). Caregivers thought that concealing a child’s HIV status would influence treatment adherence by resulting in skipped medicine (40.4%), discreet medication consumption (47.3%), and a variety of other difficulties in adhering to ART.

Furthermore, caregivers stated that adherence to ART medication encourages disclosure. When children become healthy, take medication properly, test monthly and annually, and exhibit other signs of adherence, their HIV disclosure status increases.

### Psychosocial support

Out of all participants, 258 children (68.7%) received various types of support from different organizations. Financial support was provided to 99 children (26.7%), while educational support was given to 100 children (27%). Approximately half of the caregivers (181, or 48.8%) believed that their child would face stigma due to their HIV-positive status (Table 5).

**Table 5:** Bivariable analysis result for factors associated with disclosure HIV positive status to children living with HIV in Wolaita and Hadiya zone, 2022.

| Independent Variables | Category | Disclosure status | COR (95% CI) | P value |
| --- | --- | --- | --- | --- |

|  |  | **Not disclosed** | **Disclosed** |  |  |
| --- | --- | --- | --- | --- | --- |
| Type of religion | Protestant | 120(60.3%) | 79(39.7%) | 0.370[0.156, 0.879] | 0.024** |
|  | Orthodox | 87(59.2%) | 60(40.8%) | 0.388[0.161, 0.936] | 0.035** |
|  | Muslim | 9(36%) | 16(64%) | 1 |  |
| Educational status of the caregiver | No formal education | 95(56.9%) | 72(43.1%) | 1 |  |
|  | Elementary | 78(59.1%) | 54(40.9%) | 2.122[0.731, 6.162] | 0.167 |
|  | Secondary | 29(54.7%) | 24(45.3%) | 1.938[0.659, 5.699] | 0.229 |
|  | Diploma and above | 14(73.7%) | 5(26.3%) | 2.317[0.730, 7.359] | 0.154 |
| Pediatrics duration on ART(year) | <5 | 139(54.7%) | 115(45.3%) | 1 |  |
|  | 6-15 | 77(65.8%) | 40(34.2%) | 1.593[1.011, 2.510] | 0.045** |
| Pediatrics responsible for ART | Yes | 150(58.8%) | 105(41.2%) | 0.924[0.593,1.441] | 0.727 |
|  | No | 66(56.9%) | 50(43.1%) | 1 |  |
| Caregiver discussion with a healthcare provider about disclosure | Yes | 24(42.1%) | 33(57.9%) | 2.164[1.220, 3.837] | 0.008** |
|  | No | 192(61.1%) | 122(38.9%) | 1 |  |
| pediatrics gets stigmatized | Yes | 88(48.6%) | 93(51.4%) | 1 |  |
|  | No | 128(67.4%) | 62(32.6%) | 2.182[1.433, 3.323] | 0.001** |

\*\* - Made to show statistically significant association. 1- Is representing referent

### Factors that affect the HIV-positive status disclosure for children living with HIV in Wolaita and Hadiya zone

The bivariate analysis result showed that the type of religion was significantly associated with HIV-positive status disclosure for children living with HIV. Additionally, educational status, age of pediatrics, caregiver fear of pediatric stigma and discrimination, caregiver discussion with the health care provider on disclosure, and child duration on ART were also found to be significantly associated (Table 6).

**Table 6:** Multivariable analysis result for factors associated with disclosure HIV positive status to children living with HIV in Wolaita and Hadiya zone, 2022.

| **Independent Variables** | **Category** | **Disclosure status** |  | **AOR (95% CI)** | **P – value** |
| --- | --- | --- | --- | --- | --- |
|  |  | **Not disclosed** | **Disclosed** |  |  |
| Child duration on ART | <5 | 139(54.7%) | 115(45.3%) | 1 |  |
|  | 6-15 | 77(65.8%) | 40(34.2%) | 1.633[1.013, 2.631] | 0.044** |
| Type of religion | Protestant | 120(60.3%) | 79(39.7%) | 0.462[0.187, 1.138] | 0.093 |
|  | Orthodox | 87(59.2%) | 60(40.8%) | 0.446[0.179, 1.116] | 0.084 |
|  | Muslim | 9(36%) | 16(64%) | 1 |  |
| Caregiver discussion with healthcare providers about disclosure | Yes | 24(42.1%) | 33(57.9%) | 2.171[1.199, 3.931] | 0.011** |
|  | No | 192(61.1%) | 122(38.9%) | 1 |  |
| Educational status of the caregiver | No formal education | 95(56.9%) | 72(43.1%) | 1 |  |
|  | Elementary | 78(59.1%) | 54(40.9%) | 1.691[0.569, 5.028] | 0.334 |
|  | Secondary | 29(54.7%) | 24(45.3%) | 1.625[0.538, 4.908] | 0.389 |
|  | Diploma and above | 14(73.7%) | 5(26.3%) | 1.861[0.572, 6.058] | 0.302 |
| Caregiver fear of child getting stigmatized | Yes | 88(48.6%) | 93(51.4%) | 1 |  |
|  | No | 128(67.4%) | 62(32.6%) | 2.103[1.361, 3.250] | 0.001** |

\*\* - Made to show statistically significant association. 1 - Is representing referent

Variables that were significantly associated in the bivariable analysis, along with those having a p-value of 0.25, were selected as candidates for the multivariable analysis. Child duration on ART, caregiver discussions about disclosure with the health care provider, and caregiver fear of child stigma and discrimination were all found to be significantly associated with disclosure status among the variables considered in the multivariable analysis (Table 7).

## Discussion

Sharing HIV status with children can help prevent HIV transmission, increase drug adherence, encourage self-care, improve psychological preparedness, and improve long-term health and well-being in pediatric patients. However, shame and fear frequently cause caregivers to conceal their HIV status from children. The purpose of this study was to determine the extent to which pediatric HIV-positive children disclose their status and to investigate the factors that influence disclosure in the Wolaita and Hadiya zones.

In this study, the overall pediatric HIV-positive status disclosure rate was 41.8% (95% CI = 37– 47). This finding is consistent with the findings of a study conducted in the North Gondar zone, which was 39.5%; the North Shoa zone, which was 43.6%; and the Bench Sheko and West Omo zones, which were 45.6%. This consistency in findings may be attributed to similarities in the study results rather than the study population and data collection method (15-17). This finding is higher than that of studies conducted in South India, which was 10.3%; Ghana, which was 33.3%; Tanzania, which was 33%); Bahir Dar, which was 31.5%; Bale Zone, which was 28.5%; and East Gojjam, which was 33.3%. The potential variation could be attributed to changes in the study period, as community awareness about HIV and related information improves over time, potentially leading to increased disclosure by caregivers. Moreover, socio-cultural differences may contribute to the disparity between these studies (18-23). This finding is lower than that of a study conducted in the United States, Canada, Europe, Uganda, Western Kenya, East Arsi Zone, and Dire Dawa (2, 4, 6, 24). This difference in disclosure rates may be attributed to various factors: the difference in sample size, caregivers’ fear of stigma and discrimination for their pediatric patients by the community, individual income variation, and the lack of knowledge and skills of caregivers on how to approach or inform their pediatric patients living with HIV. Additionally, the low rate of caregivers attending psychosocial support groups in this study may also contribute to the lower disclosure rate.

The findings from this study showed that caregiver discussions with health care providers about disclosure status, child duration on ART, and caregiver fear of child stigma and discrimination were significantly associated with disclosing a pediatric HIV-positive status. Caregivers who discussed the pediatric HIV-positive status with health care providers were more inclined to disclose it compared to those who did not. This result aligns with a study conducted in the Bench Sheko and West Omo zones, indicating a similar trend (17). This similarity may be attributed to commonalities in the study population and data collection methods. In contrast, this result differs from the study conducted in Dire Dawa (4). These disparities in results may stem from differences in the study period or sample size.

The length of time children spent on ART was associated with pediatric HIV-positive status disclosure. According to this finding, parents were more likely to disclose their children’s HIV-positive status if the children had ART follow-ups between 6 and 15 years old compared to those with ART follow-ups at 5 years old. This finding is similar to that of a study conducted in Bahir Dar, the East Arsi Zone, Bench Sheko, and the West Omo Zone (17, 21, 24). One possible explanation is that children who have been on ART follow-up for a long time have had opportunities to ask questions about their HIV medications and why they are taking the medication. This may empower caregivers to disclose the child’s HIV-positive status. In contrast, this finding contradicts the results from a study conducted in South Africa (25).

Caregivers’ fear of their child being stigmatized was associated with the disclosure of pediatric HIV-positive status. This study found that caregivers did not perceive children as stigmatized, and disclosure of pediatric HIV-positive status was more common than caregivers fearing stigma and discrimination for their child. This finding is consistent with a study conducted in Dire Dawa, which also reported similar results (4). One possible reason for this is that since most children with HIV contracted it from their mothers, revealing a child’s HIV-positive status may also reveal other family secrets, resulting in stigma and discrimination. Although this study did not find a link between the appropriate age for disclosure and pediatric HIV-positive status disclosure, other studies have shown that the suitable age for disclosure is twelve years and older. For example, a study in East Arsi Zone, Ethiopia, found that child age was a key factor in pediatric HIV-positive status disclosure (24). The average age for disclosure in this study was 11.88 (SD 2.4), consistent with findings in the same region. One possible reason is that as children grow older, they are more likely to ask about their illness. Consequently, caregivers can then explain the diagnosis and current health status to their children.

In addition, 31.1% of pediatric psychosocial support group participants in hospitals were found in this study. Although not significantly associated with disclosing pediatric HIV-positive status in this study, other research has shown a significant link to pediatric HIV-positive status disclosure. For example, a Ugandan study found that psychosocial support was significantly associated with pediatric HIV-positive status disclosure (2). This study found that psychosocial support group participants have low attendance rates. Caregivers who do not participate in these groups may be less likely to share their experiences, lessons learned, and HIV information with children.

### Strength and Limitation of the study

The study included a representative sample of children receiving HIV treatment at health facilities, thus the findings are applicable to comparable contexts. Desirability bias may have occurred, in which individuals fraudulently reported their conduct to conform to society standards. Furthermore, recall bias might exist because caregivers may have difficulties recalling the child’s age upon diagnosis or the length of ART. The cross-sectional research design makes it difficult to determine a definite cause-and-effect link between the variables.

## Conclusion

In this study, the prevalence of HIV-positive status declaration to HIV pediatricians is low when compared to previous Ethiopian study findings. The main reasons for the low pediatric HIV-positive disclosure status in this study were caregivers with a low level of education, a currently living child living with non-biological parents, a child who has lost any family members to HIV, a child with little responsibility for ART, a lack of caregiver discussion with a health care provider about pediatric HIV positive disclosure status, and poor caregiver attendance at a psychosocial support group. Factors linked with pediatric HIV-positive disclosure status include caregiver interactions with health care professionals concerning disclosure, child time on ART, and caregivers’ fear that their kid would be ostracized owing to their HIV-positive status. Therefore, based on the findings of this study, it is imperative to make concerted efforts to enhance pediatric HIV-positive disclosure status.

## Data Availability

We have the data used for the analysis and will make it available upon request.

## Abbreviation/Acronym

AAP: American Academy of Pediatrics
ANECCA: African Network for the Care of Children
CLHIV: Children living with HIV/AIDS
LMIC: Low and Middle-Income Countries
MTCT: Mother-to-Child Transmission

## Informed consent for study participants

Ethical procedures were followed to conduct research after ensuring that caregivers of the children were informed by the researchers. After explaining the study’s purpose, rights to participate, withdraw, confidentiality, lack of incentives, benefits, and confidentiality, contact the appropriate research oversight body to ask questions. All volunteers were recruited for the study after signing up to participate.

## Data and Software Availability

We have the data used for the analysis and will make it available upon request.

## Author Contributions

a. Conceptualization, writing – original draft preparation, data curation, formal analysis, investigation, methodology, project administration, resources, software,
b. Methodology, project administration, resources, software, supervision, validation, visualization, writing – review & editing,
c. Methodology, project administration, resources, software, supervision, validation, visualization, writing – review & editing,
d. Validation, visualization, writing – review & editing, prepared the manuscript

## Acknowledgment

I would like to express my deepest gratefulness to Wolaita Sodo University College of Medicine and Health Science for allowing me to develop this thesis. I am grateful to the Health office of Wolaita Sodo Health Administration and Hadiya zone health administration for providing me with the necessary information essential for developing the thesis. Finally, I express my deep gratitude to the study participants and everyone who assisted in completing the research, including Mr. Tesfaye Langana, Mr. Cherent Tesfaye, Mr. Zerhun Zana, and Sr. Saba Zekariyas. Their contributions in translation to the local language and encouragement are highly appreciated.

## References

1. Tucho WA, Tekelehaimanot AN, Habte MB. Disclosure Status and Associated Factors Among Children on Antiretroviral Therapy in Ethiopia. Pediatric Health Med Ther. 2021;12:299–306.

2. Namasopo-Oleja SM, Bagenda D, Ekirapa-Kiracho E. Factors affecting disclosure of serostatus to children attending Jinja Hospital Paediatric HIV clinic, Uganda. African health sciences. 2015;15(2):344–51.

3. Tadesse BT, Foster BA, Berhan Y. Cross Sectional Characterization of Factors Associated with Pediatric HIV Status Disclosure in Southern Ethiopia. PLoS One. 2015;10(7):e0132691.

4. Guta A, Areri HA, Anteab K, Abera L, Umer A. HIV-positive status disclosure and associated factors among children in public health facilities in Dire Dawa, Eastern Ethiopia: A cross-sectional study. PLoS One. 2020;15(10):e0239767.

5. Britto C, Mehta K, Thomas R, Shet A. Prevalence and Correlates of HIV Disclosure Among Children and Adolescents in Low- and Middle-Income Countries: A Systematic Review. J Dev Behav Pediatr. 2016;37(6):496–505.

6. Turissini ML, Nyandiko WM, Ayaya SO, Marete I, Mwangi A, Chemboi V, et al. The prevalence of disclosure of HIV status to HIV-infected children in Western Kenya. Journal of the Pediatric Infectious Diseases Society. 2013;2(2):136–43.

7. Vaz LM, Maman S, Eng E, Barbarin OA, Tshikandu T, Behets F. Patterns of disclosure of HIV status to infected children in a Sub-Saharan African setting. J Dev Behav Pediatr. 2011;32(4):307–15.

8. Balcha Berhanu AA. Challenges of Caregivers to Disclose their Children’s HIV Positive Status Receiving Highly Active Anti Retroviral Therapy at Pediatric AntiRetroviral Therapy Clinics in Bahir Dar, North West Ethiopia. Journal of AIDS & Clinical Research. 2013;04(01).

9. Lencha B, Ameya G, Minda Z, Lamessa F, Darega J. Human immunodeficiency virus infection disclosure status to infected school aged children and associated factors in bale zone, Southeast Ethiopia: cross sectional study. BMC Pediatr. 2018;18(1):356.

10. Biadgilign S, Deribew A, Amberbir A, Escudero HR, Deribe K. Factors associated with HIV/AIDS diagnostic disclosure to HIV infected children receiving HAART: a multi-center study in Addis Ababa, Ethiopia. PLoS One. 2011;6(3):e17572.

11. Federal ministry of Ethiopia. National guidelines for comprehensive HIV prevention, care and treatment. Addis Ababa, Ethiopia,2017.

12. Tucho WA, Tekelehaimanot AN, Habte MB. Disclosure status and associated factors among children on antiretroviral therapy in Ethiopia. Pediatric Health, Medicine and Therapeutics. 2021:299–306.

13. Sariah A, Rugemalila J, Somba M, Minja A, Makuchilo M, Tarimo E, et al. “Experiences with disclosure of HIV-positive status to the infected child”: Perspectives of healthcare providers in Dar es Salaam, Tanzania. BMC Public Health. 2016;16(1):1083.

14. Berhane Alema H. HIV Positive Status Disclosure and Associated Factors among HIV Positive Adults in Axum Health Facilities, Tigray, Northern Ethiopia. Science Journal of Public Health. 2015;3(1):61.

15. Negese D, Addis K, Awoke A, Birhanu Z, Muluye D, Yifru S, et al. HIV-positive status disclosure and associated factors among children in North Gondar, Northwest Ethiopia. International Scholarly Research Notices. 2012;2012.

16. Shallo SA, Tassew M. HIV Status Disclosure and its associated factors among Children on Antiretroviral Therapy in West Shoa Zone, Ethiopia, 2019: A Mixed method cross-sectional study. 2019.

17. Tucho WA, Tekelehaimanot AN, Habte MB. Disclosure Status and Associated Factors Among Children on Antiretroviral Therapy in Ethiopia. Pediatric Health, Medicine and Therapeutics. 2021;12:299.

18. Ekstrand ML, Heylen E, Mehta K, Sanjeeva G, Shet A. Disclosure of HIV status to infected children in South India: perspectives of caregivers. Journal of tropical pediatrics. 2018;64(4):342–7.

19. Gyamfi E, Okyere P, Enoch A, Appiah-Brempong E. Prevalence of, and barriers to the disclosure of HIV status to infected children and adolescents in a district of Ghana. BMC international health and human rights. 2017;17(1):1–8.

20. Nzota MS, Matovu JK, Draper HR, Kisa R, Kiwanuka SN. Determinants and processes of HIV status disclosure to HIV-infected children aged 4 to 17 years receiving HIV care services at Baylor College of Medicine Children’s Foundation Tanzania, Centre of Excellence (COE) in Mbeya: a cross-sectional study. BMC pediatrics. 2015;15(1):1–9.

21. Alemu A, Berhanu B, Emishaw S. Challenges of caregivers to disclose their children’s HIV positive status receiving highly active antiretroviral therapy at pediatric antiretroviral therapy clinics in Bahir Dar, North West Ethiopia. J AIDS Clin Res. 2013;4(253):1–7.

22. Lencha B, Ameya G, Minda Z, Lamessa F, Darega J. Human immunodeficiency virus infection disclosure status to infected school aged children and associated factors in bale zone, Southeast Ethiopia: cross sectional study. BMC pediatrics. 2018;18(1):1–8.

23. Tamir Y, Aychiluhem M, Jara D. Disclosure Status and Associated Factors among Children Living With HIV in East Gojjam, Northwest of Ethiopia 2014. Quality in Primary Care. 2015;23(4).

24. Yami DB, Tuji TS, Gelete BWM, Beyene Workie K. Disclosure status of HIV-positive children and associated factors among children in public health facilities in East Arsi zone, Oromia regional state, South Eastern Ethiopia: A cross-sectional study. SAGE open medicine. 2022;10:20503121211068725.

25. Madiba S. Patterns of HIV diagnosis disclosure to infected children and family members: data from a paediatric antiretroviral program in South Africa. World Journal of AIDS. 2012;2(03):212.

